# Stem Cell-Derived Gene Expression Scores Predict Survival and Blastic Transformation in Myelofibrosis

**DOI:** 10.1101/2024.07.09.24310101

**Authors:** Jessie J.F. Medeiros, Andy G.X. Zeng, Michelle Chan-Seng-Yue, Tristan Woo, Suraj Bansal, Hyerin Kim, Jessica L. McLeod, Andrea Arruda, Hubert Tsui, Jaime O. Claudio, Dawn Maze, Hassan Sibai, Mark D. Minden, James A. Kennedy, Jean C.Y. Wang, John E. Dick, Vikas Gupta

## Abstract

**Purpose:** Myelofibrosis (MF) is the most severe myeloproliferative neoplasm (MPN) where there remains a need for improved risk stratification methods to better inform patient management. Since MF is a stem cell driven disease and stem cell informed transcriptomic information has been shown to be prognostic across other clinical settings we sought to use this information to generate novel transcriptomic-based risk stratification models that could complement current approaches.

**Patients and Methods:** We identified 358 MF patients from the MPN registry at the Princess Margaret Cancer Centre (ClinicalTrials.gov Identifier: NCT02760238) from whom peripheral blood mononuclear cells were collected and clinical data was available. We randomly split our cohort into a 250-patient training set and a 108-patient test set to train and validate prognostic models, respectively.

**Results:** Within the training set we used repeated nested cross validation together with LASSO regression from various starting gene sets and found that the best prognostic models were consistently derived from transcriptomic variation among MF stem cells. From this gene set we trained our final model, a 24-gene weighted expression score (termed, MPN24) that is prognostic for overall survival. Patients were classified as MPN24-High or MPN24-Low risk depending on whether their scores were above or below the within cohort median defined in the training set. The prognostic power of MPN24 was validated in the test set patients with stark differences in survival outcomes for MPN24-High (5-year survival rate = 21% [95% CI 9%-52%]) and MPN24-Low risk patients (5-year survival rate = 71% [95% CI 57-88%]) patients, resulting in a HR of 5.3 (95% CI: 2.6-10.5; p=2.08e-6). MPN24 captures unique prognostic information to current risk stratification models such as DIPSS, MIPSS70 and the Genomic-Personalized Risk scores. Therefore, we present a novel 3-tier risk stratification approach that integrates DIPSS and MPN24 to more effectively risk stratify MF patients, particularly via up or downscaling patient risk within the DIPSS-Intermediate-1/2 categories. In this integrated model patients were classified as Integrated-Low, Integrated-Intermediate or Integrated-High, and experienced 5-year survival rates of 88.2% [95% CI 77.9% - 99.9%], 39.3% [95% CI 19.9% - 77.7%], and 10.8% [95% CI 2.1% - 55.8%], respectively (likelihood ratio test p = 1e-8). Finally, from MPN24 genes we derived a 13-gene subsignature (termed, MPN13) from the training set patients that was validated to predict time-to-transformation in the test set patients when classified as MPN13-High or Low relative to the 80th percentile of MPN13 scores from the training set (p=0.0047). In the test set, MPN13-High and MPN13-Low patients experienced 3-year cumulative incidences of transformation of 5.2% [95% CI 0.2%-10.2%] and 28.6% [95% CI 3.1%-54.0%] respectively, after adjusting for death as a competing risk.

**Conclusions:** Transcriptomic information informed by MF stem cells offer novel and unique prognostic potential in MF that significantly complements current approaches. Future work will be needed to validate the robustness of the approach in external cohorts and identify how patient management can be optimized with these novel transcriptomic biomarkers.

## INTRODUCTION

Primary Myelofibrosis (PMF) is a myeloproliferative neoplasm (MPN) that originates at the level of the hematopoietic stem cell (HSC). Patients with polycythemia vera (PV) and essential thrombocythemia (ET) can experience progression of disease to myelofibrosis (post-PV MF and post-ET MF, respectively), and upon progression, have clinical features and disease course similar to PMF (Sangle et al. 2014; Masarova et al. 2017). These disease entities are collectively referred to as MF in this report. Patients with MF experience vastly different clinical outcomes, with survival ranging from months to years and variable risk of transformation to blast phase (BP) at which point survival outcomes are dismal (Kennedy et al. 2013). Allogeneic hematopoietic cell transplantation (HCT) is the only potentially curative approach, but is associated with high treatment-related morbidity and mortality (Gupta, Hari, and Hoffman 2012; Kröger et al. 2024). Meanwhile, wider availability of JAK inhibitor (JAKi) therapy, such as Ruxolitinib, has become standard of care but is limited to symptomatic control and may become inadequate for some patients. In this context, the optimal timing of HCT is not yet well defined and selection of upfront HCT versus transplant after the failure of JAKi therapy remains challenging (Maze et al. 2024; Rajendra and Gupta, Current Heme Malignancy Reports, 2024, paper under review). Accurate prognostication is key to guiding patient care in this clinically heterogeneous group.

Current risk prediction models such as the DIPSS score use clinical and laboratory features such as age and blood counts to predict MF patient risk and help guide clinical decision-making (Passamonti et al. 2010). More recently, MIPSS70, MIPSS70-plus and personalized risk scores (Genomic-PRS) have additionally incorporated mutation and/or cytogenetics data to augment risk stratification (Guglielmelli et al. 2018; Grinfeld et al. 2018). These tools highlight the added prognostic value of incorporating molecular information pertaining to genomics. Yet, existing prognostic models in MF lack transcriptomic information, which has been shown to be highly prognostic in other cancers (Nielsen et al. 2010).

Furthermore, features of disease-driving stem cells have been used successfully to derive prognostic scores in other blood cancers (Ng et al. 2016; Gentles et al. 2010). Thus, we hypothesized that transcriptomic variation among disease-driving MF stem cells may provide novel prognostic information independent from the genomic and clinical features used in present-day clinical practice. Further, a biomarker that captures this variation may complement and improve existing approaches for prognostic stratification of MF.

Here we present a large-scale transcriptomic cohort of 358 MF patients linked to clinical outcomes captured in a prospective registry. We leverage transcriptomic variation corresponding to both intra- and inter-patient heterogeneity among MF stem cells to train and validate, a novel 24-gene weighted expression score (termed, MPN24) predictive of survival and a 13-gene subscore (termed, MPN13) predictive of blastic transformation in MF. Critically, MPN24 is independently prognostic after accounting for genomic and clinical features and complements present-day risk stratification approaches in MF. Improved identification of high-risk MF patients enabled by MPN24 may facilitate clinical decision-making around optimal timing of HCT and prioritization of patients for clinical trials.

## PATIENTS AND METHODS

### Patient Cohort

A total of 358 patients included in this study were identified from the MPN registry at the Princess Margaret Cancer Centre (ClinicalTrials.gov Identifier: NCT02760238) from whom peripheral blood mononuclear cells (PB-MNCs) were collected at the first visit to Princess Margaret. Inclusion criteria consisted of a diagnosis of MF including PMF, pre-fibrotic MF, post-ET MF or post-PV MF in chronic phase. Individuals were excluded if they received a diagnosis of MPN-Unclassified (MPN-U), MPN/myelodysplastic syndrome (MDS) overlap or lacked a biospecimen. All biological samples were collected under Research Ethics Board approval (REB#17-5601) with written informed consent as per the Declaration of Helsinki and viably frozen in the Princess Margaret, hematologic malignancies tissue bank.

### Sample preparation and RNA sequencing

To prepare samples for RNAseq cells were thawed dropwise, centrifuged and resuspended in 1 mL of PBS 5% FCS with 100X DNase1. Up to 2 million unsorted viable cells were aliquoted into RNA Lysis Buffer (RLT; RNeasy Minikit; Qiagen). RLT samples were stored at -80C and RNA was later extracted using the RNeasy Minikit (Qiagen) following the “Quick-Start Protocol” with “On-Column DNase Digestion”. RNA was eluted in 30ul of molecular-grade water and quantified using the High-Sensitivity RNA Qubit Assay (Invitrogen). RNA samples were run on the High-Sensitivity BioAnalyzer (Agilent) to determine RNA quality. RNA samples with an RNA integrity number (RIN) > 8 proceeded to PolyA enrichment and directional library preparation at The Center for Applied Genomics (SickKids, Toronto, Canada). Libraries were sequenced on the NovaSeq S4 at a read-length of 100bp, targeting approximately 50 million paired-end reads per sample.

### RNAseq alignment, transcript count data and normalization

RNA-seq data was aligned to GRCh38 with STAR v2.7.9a using the two-pass mode. The following parameters were set: chimSegmentMin=12, chimJunctionOverhangMin=8, alignSJDBoverhangMin=10, alignIntronMax=100000, alignSJstitchMismatchNmax=5 -1 5 5, alignSplicedMateMapLminOverLmate=0, alignSplicedMateMapLmin=30, chimMultimapScoreRange=3, chimScoreJunctionNonGTAG=-4, chimMultimapNmax=20, chimNonchimScoreDropMin=10 peOverlapNbasesMin=12 and peOverlapMMp=0.1. Reads were annotated with Ensembl v103. Count data was obtained using HTSeq v0.7.2. General stats were obtained using picard v2.10.9.

The RNAseq data was processed through DESeq2 v1.32.0. Genes with less than 1 read were removed prior to normalization using variance stabilized transformation (VST).

### Machine learning approach for prognostic model generation

To train and then validate novel prognostic scores in MF, we randomly split our cohort of PB-MNCs (n=358) into train (70%; n =250) and test (30%; n=108) sets, respectively. Importantly, we confirmed that there were no statistically significant differences between the train and test sets across 25 clinically relevant parameters. To train gene expression scores for predicting patient survival using a minimal set of features/genes, we employed Cox proportional hazards regression using least absolute shrinkage and selection operator (LASSO) on data generated only in the train set (n = 250). Further to this approach, we used repeated nested cross validation to mitigate the risk of overfitting models to the train set. LASSO regression together with repeated-nested cross validation generates a distribution of p-values for a given model configuration (i.e. the specific starting genes inputted into the model, the regression approach used and the data associated with samples included in the training set) such that the configuration that consistently generates the best models can be identified and selected for use in the generation of a final prognostic model.

### Survival analysis

Survival analysis was performed using the *survival, survminer* packages in R. Overall survival (OS) outcomes were censored at the time of transplant in order to focus on MF-related outcomes independent of transplant. Kaplan-meier plots were used to visualize differences in survival outcomes between groups of patients. Cox proportional hazards regression was used to quantify association with survival for both univariable and multivariable analysis. To assess for independent prognostic value of a biomarker in multivariable analysis, a nested likelihood ratio test was performed to quantify significance in model improvement after biomarker incorporation, relative to a baseline model composed only of the covariates. For analyses pertaining to time-to-transformation, competing risk analysis was performed using the *cmprsk* R package.

## RESULTS

### Variation among MF stem and progenitor cells enable the development of highly prognostic models for prediction of survival in myelofibrosis

Our experimental approach for prognostic model generation is summarized in **Figure 1**. Briefly, to identify the model configuration (i.e. starting gene set and LASSO penalty value) that consistently produced the best models for prediction of survival outcomes in MF, we applied LASSO regression together with repeated nested cross-validation on the 250-patient training set. Performance of prognostic models depends heavily on the features selected for model training. Thus, we trained models using different sets of features ranging from the whole transcriptome to smaller sets of biologically motivated genes. These biologically motivated genesets spanned broad categories including, leukemia stem cell (LSC) genes from AML patients (Ng et al. 2016), variable genes across our MF cohort, and variable genes from CD34+ MF stem and progenitor cells (Psaila et al. 2020). For each starting set of features, our cross-validation approach trained 500 prognostic models on random subsets of the data and produces a distribution of p-values depicting multivariable prognostic significance adjusted for clinically relevant features (i.e. DIPSS score, ECOG status, PB blast % and age) for each model configuration. This allowed us to identify the starting features which most robustly produced the best performing prognostic models within our data. Critically, the most accurate prognostic models were consistently produced from highly variable genes derived from scRNA-seq profiles of 82,255 Lin-CD34+ MF stem and progenitor cells across 15 patients (Psaila et al. 2020). This represents a significant improvement over model training from the entire transcriptome or LSC genesets defined from AML samples. Thus, features of intra- and inter-patient heterogeneity specific to MF stem and progenitor cells proved to be most relevant for predicting survival.

**Figure 1.**
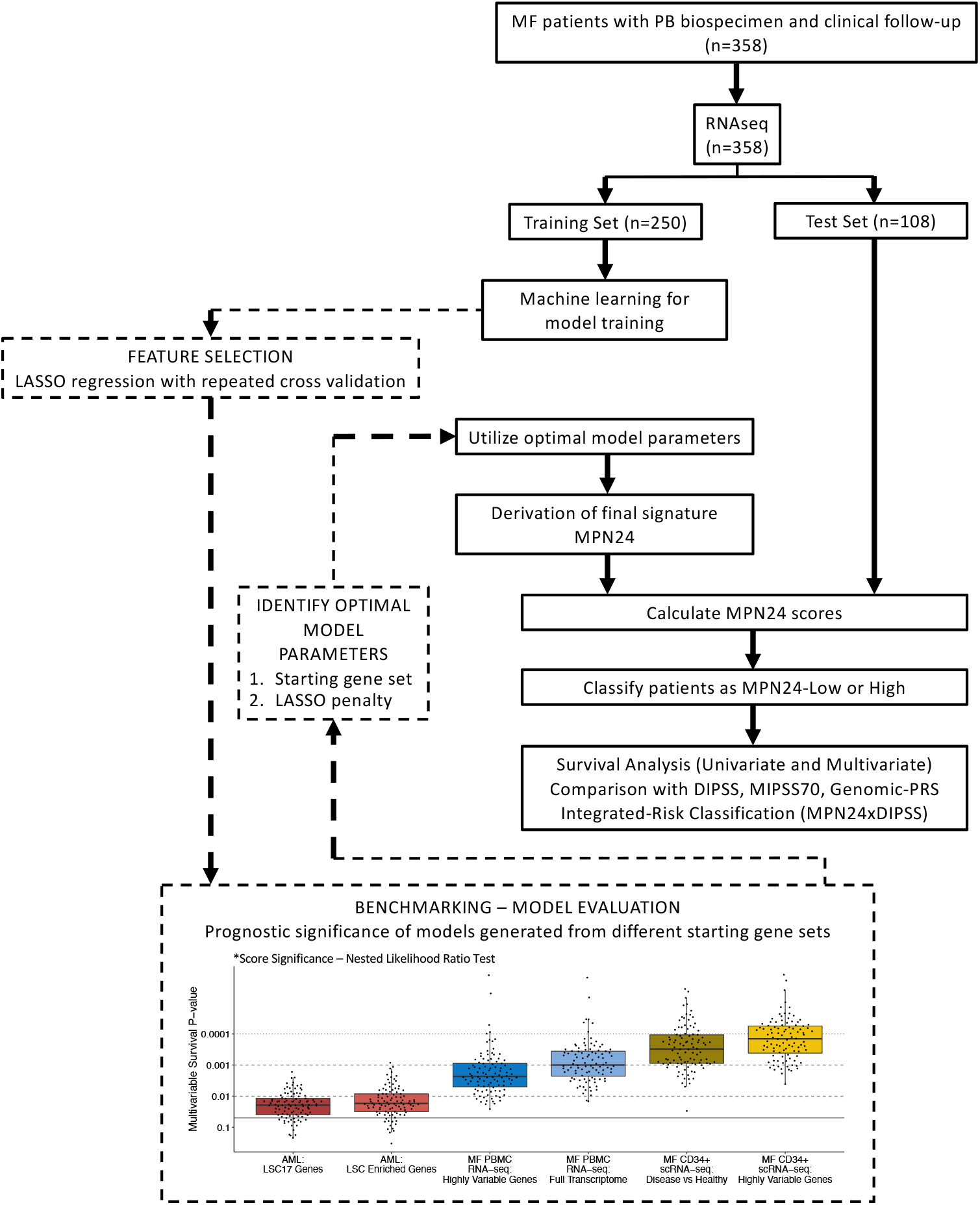
Machine learning approach for derivation and validation of a 24-gene expression score predictive of survival in MF. CONSORT diagram summarizing study design. Our PMCC-MF cohort (n=358) was split into a 250-patient training set and a 108-patient test set. Within the 250-patient training set, our machine learning approach utilized LASSO regression together with repeated nested cross validation to identify the most optimal model parameters (i.e. gene set and LASSO penalty value) on which to train our final model from. For example, models were trained from the full transcriptomic dataset generated in our study (i.e. “MF PBMC RNA-seq: Full Transcriptome”) as well as more condensed biologically motivated gene sets including AML LSC-related genes (i.e. “AML: LSC17 genes” and “AML: LSC Enriched Genes”; Ng et al., 2016), variable genes from our RNAseq cohort (i.e. “MF PBMC RNA-seq: Highly Variable Genes” and MF CD34+ genes derived from our retrospective analysis of published scRNAseq of MF CD34+ cells (i.e. “MF CD34+ scRNA-seq: Disease vs Healthy” and “MF CD34+ scRNA-seq: Highly Variable Genes”; Psaila et al., 2020). In benchmarking analysis for model evaluation, the MF CD34+ gene set capturing the most highly variable genes among MF stem cells (i.e. “MF CD34+ scRNA-seq: Highly Variable Genes”) consistently produced significantly better models than those derived from other gene sets, including the entire transcriptome. Therefore, this gene set and an optimal lambda penalty value were used with LASSO on the full training set (n=250) to train the final prognostic signature. Our final model, termed MPN24, consists of 24 genes whose weighted sum of expression gives rise to a prognostic signature for OS in MF. MPN24 scores were then calculated from the RNAseq data in all train and test set patients. The within cohort median split from the training set was used to classify all patients as either MPN24-High or MPN24-Low if their scores were above or below this threshold, respectively. These designations were used for all subsequent survival analysis including integration analyses with currently used risk stratification approaches such as DIPSS, MIPSS70 and PRS. *P value from Nested Likelihood Ratio Test to evaluate model improvement compared against a baseline model of DIPSS, ECOG, PB blast % and age.

Since the scRNAseq data generated from MF CD34+ stem cells most robustly give rise to the best prognostic models in MF we used all 250 patients in our train set to derive the final model using this starting geneset. This approach generated a final model calculated as the weighted sum of gene expression across 24 genes (termed, MPN24). MPN24 scores were then calculated for all samples in the train and test sets.

### MPN24 is a stem cell derived gene expression signature that predicts overall survival in myelofibrosis

To evaluate the prognostic potential of MPN24 to predict OS, we first dichotomized patients with MPN24 scores above or below the training cohort median as MPN24 high or low, respectively. Within the training set, we observed stark differences in survival outcomes between MPN24-High (5-year survival rate = 21% [95% CI: 13%-32%]) and MPN24-Low (5-year survival rate = 85% [95% confidence interval (CI): 77-94%]) patients, resulting in a hazard ratio (HR) of 10.9 (95% CI: 6.3-18.6; p < 2e-16) (**Figure 2a**).

**Figure 2.**
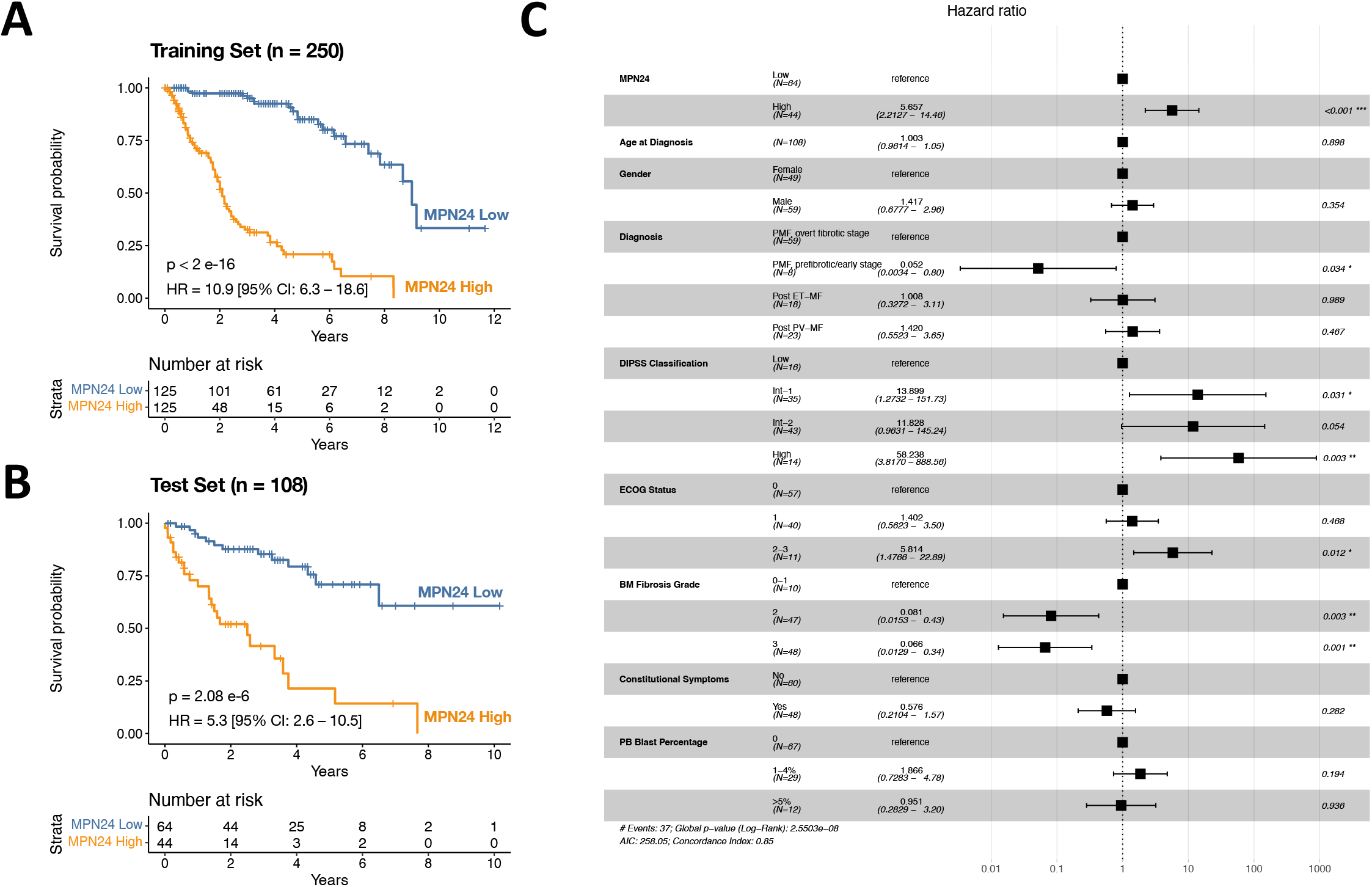
MPN24 as a binary test predicts OS in MF patients. MPN24 is prognostic for OS when used as a binary test to classify patients as MPN24-low or MPN24-high scoring in relation to their scores being below or above the MPN24 training set median, respectively. **a**. Kaplan-Meier plot displaying MPN24 score performance in the training set (n=250) that was used to generate the model. **b**. Kaplan-Meier plot displaying validation of MPN24 score performance in the test set (n=108) left out of signature training. **c**. Forest plot displaying multivariate analysis of MPN24 binary classification in the test set (n=108) when incorporating various clinical features. *Outcomes for all analyses are censored at the time of HCT.

Importantly, we validated these results in patients within the test set (n=108) that were excluded from model training. In the test set, significant differences in OS persisted between MPN24-High (5-year survival rate = 21% [95% CI 9%-52%]) and MPN24-Low (5-year survival rate = 71% [95% CI 57-88%]) patients, resulting in a HR of 5.3 (95% CI: 2.6-10.5; p=2.08e-6) (**Figure 2b**). Furthermore, MPN24-based patient stratification within the test set (n = 108) retained independent prognostic value in multivariate analysis incorporating age, sex, DIPSS category, ECOG status, fibrosis grade, constitutional symptoms, and PB blast percentage (**Figure 2c**).

### MPN24 captures prognostic features independent of commonly used risk stratification models such as DIPSS and MIPSS70

Since DIPSS and MIPSS70 are two of the most commonly utilized risk stratification models in clinical management of MF, we evaluated the ability of MPN24 to stratify patients within these predefined risk categories. Importantly, this analysis was performed in the test set patients (n=108) that were excluded from MPN24 training to avoid potential biases resulting from model overfitting in the training cohort. When intersecting DIPSS with MPN24 there are MPN24 high and low scoring test set patients across all DIPSS categories. In survival analysis MPN24 retains prognostic value in multivariable analysis and separates high and lower risk patients within pre-defined DIPSS categories including those classified as DIPSS Low/Int-1 (HR = 3.82; p = 0.035) and DIPSS Int-2/High (HR = 3.95; p = 0.00059) (**Figure 3a**).

**Figure 3.**
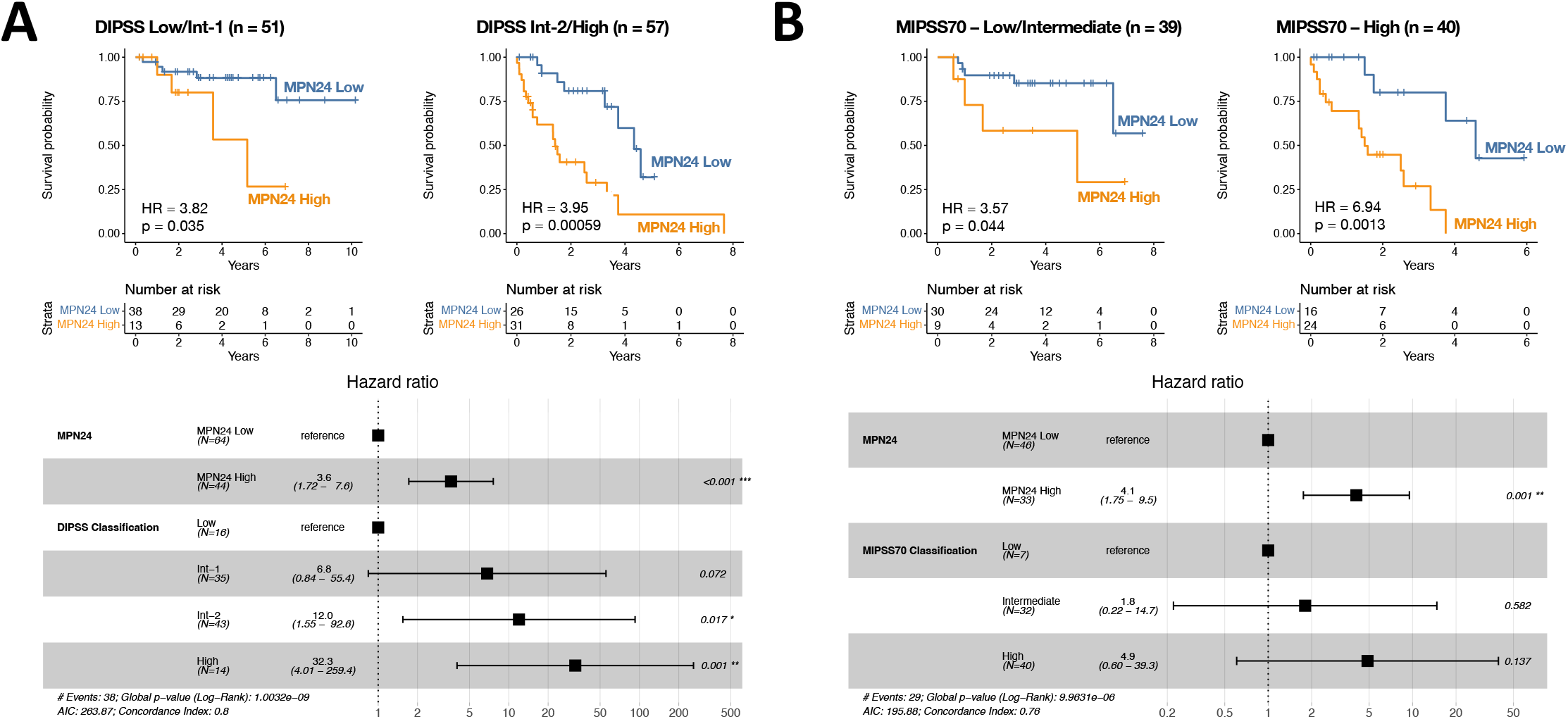
Comparing MPN24 performance in relation to classically-defined risk stratification models in test set patients (n=108). **a**. MPN24 in predefined DIPSS categories. Top panel: Kaplan-Meir plots displaying the prognostic utility of MPN24 to refine prognostication in DIPSS-Low/Intermediate-1 (left) and DIPSS Intermediate-2/High (right) categories. Bottom panel: Forest plot displaying multivariate analysis of MPN24-Low and MPN24-High against DIPSS categories. **b**. MPN24 in predefined MIPSS70 categories. Top panel: Kaplan-Meir plots displaying the prognostic utility of MPN24 to refine prognostication in MIPSS70-Low/Intermediate (left) and MIPSS70 High (right) categories. Bottom panel: Forest plot displaying multivariate analysis of MPN24-Low and MPN24-High against MIPSS70 categories. *Outcomes for all analyses are censored at the time of HCT.

Similarly, when intersecting MIPSS70 categories with MPN24 there are MPN24 high and low scoring patients in each previously defined class for test set patients. In the test set, we limited our analysis to 79 of the108 patients where we had access to mutational data required for MIPSS70 classification. Since our cohort was skewed toward MIPSS70-Intermediate and High-risk groups with only 7 patients classified as MIPSS70-Low, we combined MIPSS70-Low and MIPSS70-Intermediate patients in our analysis. In survival analysis MPN24 retains prognostic value in multivariable analysis and separates high and lower risk patients in pre-defined MIPSS70 categories, including those classified as MIPSS70-Low/Intermediate (HR = 3.57; p = 0.044) and MIPSS70-High (HR = 6.94; p = 0.0013) (**Figure 3b**). For the 79 patients with full clinical and mutational data we performed univariate analysis demonstrating the c-index increases significantly when MPN24 is added to traditional models, including DIPSS, MIPSS70 and Genomic-PRS. Therefore, MPN24 is independently prognostic and captures information unique and complementary to models informed by clinical and/or mutational data.

### A new 3-tiered risk stratification approach that integrates MPN24 with DIPSS

Within the DIPSS classification, most patients are positioned within the Intermediate categories (DIPSS-Intermediate-1 and Intermediate-2) where clinical decision making is most unclear. Given that MPN24 captures prognostic information independent of DIPSS, we asked whether DIPSS could be integrated with MPN24 to reduce the size of the intermediate risk category and confidently re-assign more patients into high-risk and low-risk categories. To this end, we compared the distribution of MPN24 and DIPSS categories within the test set patients (n=108) and generated a new 3-tiered risk stratification approach (i.e. DIPSSxMPN24-Integrated Model). First, patients classified as DIPSS-Low or DIPSS-Intermediate-1 and MPN24-Low were newly classified as “Integrated-Low” (n=38). Second, patients classified as DIPSS-Intermediate-2 or DIPSS-High and MPN24-High were newly classified as “Integrated-High” (n=31). The remaining patients (n = 39) had transcriptional features that were discordant with their clinical features, resulting in conflicting risk classifications by DIPSS and MPN24. These patients were consolidated into an “Integrated-Intermediate” category (n=39) (**Figure 4a**). This new integrated classification effectively upscaled or downscaled patient risk, including 59% of the patients (n=46/78) belonging to DIPSS-Intermediate categories (i.e. DIPSS-Intermediate-1 and DIPSS-Intermediate-2) that were reclassified as Integrated-High (n=21/78; 27%) or Integrated-Low (n=25/78; 32%). This results in a model where patients are more evenly distributed across risk categories with the size of the Intermediate category being reduced from 78 (72%) patients by DIPSS to 39 (36%) patients in the Integrated model (**Figure 4b**). Using this integrated model, patients classified as Integrated-Low, Integrated-Intermediate or Integrated-High, experienced 5-year survival rates of 88.2% [95% CI 77.9% - 99.9%], 39.3% [95% CI 19.9% - 77.7%], and 10.8% [95% CI 2.1% - 55.8%], respectively (likelihood ratio test p = 1e-8) (**Figure 4c**).

**Figure 4.**
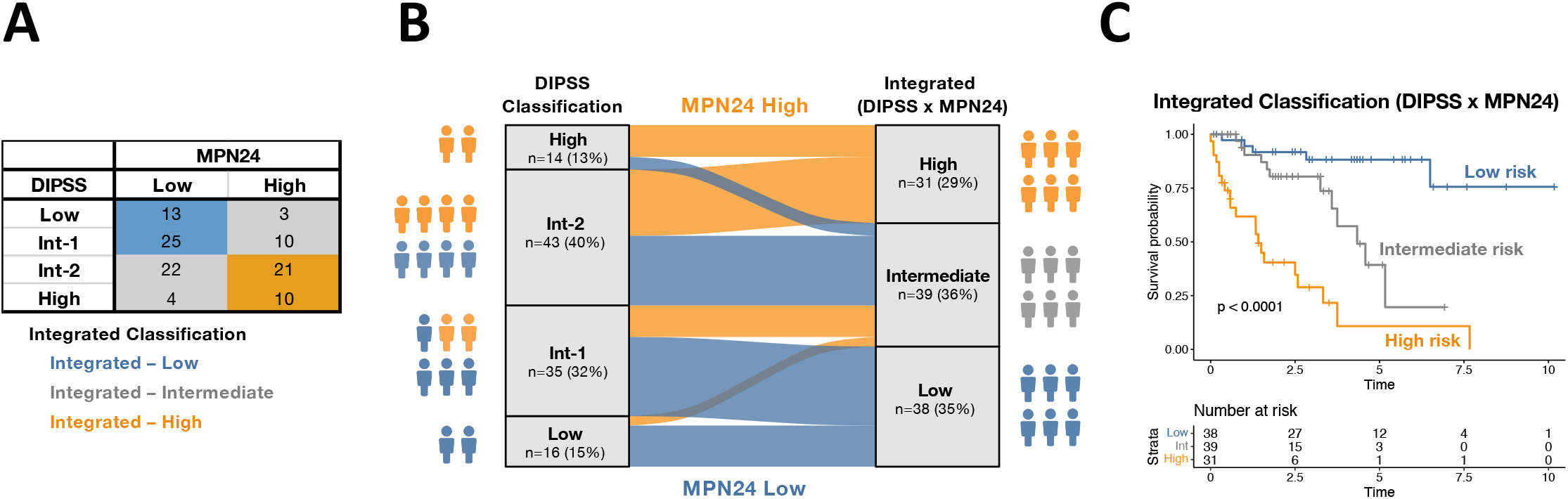
A new 3-tier risk stratification approach for MF that integrates DIPSS with MPN24. **a**. Matrix of test set patients classified by DIPSS and MPN24. Three new integrated risk categories are formed (Integrated-Low, Integrated-Intermediate and Integrated-High) based on the intersection between the DIPSS and MPN24 risk categories. **b**. Sankey diagram illustrating the number and percentage of individuals in each DIPSS category (left) that are redistributed via MPN24-High (orange) and MPN24-Low (blue) into the 3-tier integrated risk classifications (right). **c**. Kaplan Meier plots displaying the prognostic significance of the Integrated model (Low, Intermediate, High) for predicting patient survival. *Outcomes are censored at the time of HCT.

### A 13-gene MPN24 sub-signature that predicts risk of blastic transformation in MF patients

Given that blastic transformation is a negative contributor to OS we hypothesized that genes in the MPN24 score might also capture features specific to blastic transformation in MF. Therefore, we used the 24 genes from MPN24 and regressed on time-to-transformation (censoring all other events) within 5 years of sample collection to derive a new 13-gene subscore (termed, MPN13) predictive of blastic transformation by using all of the data in the training set (n=250). This 5-year cutoff was chosen to select for individuals who transform to BP relatively soon after sample collection but within a window of opportunity for intervention with, for example, HCT (Barbui et al. 2011; Vannucchi et al. 2015). Through nested cross-validation, we found that models predicting time-to-transformation trained from the original 24 genes within MPN24 led to consistently better performance in predicting time-to-transformation than models trained on other sets of features including the whole transcriptome.

To generate a binary approach to classification using MPN13, we classified patients above and below the 80th percentile from the training set data as high and low risk, respectively. We chose a higher cut-off for setting the binary classification for MPN13 in contrast to MPN24 (i.e. where the training set median threshold was used) since only a small number of individuals transformed to BP within 5 years of sample collection in our cohort (n=33/250; 13% in the training set). Despite the small number of events, binary classification using MPN13 was significantly associated with risk of transformation (p=1.1e-16) in the training set by competing risk analysis. More importantly, using the 80^th^ percentile cut-off defined in the training set data, MPN13 was also validated in the test set (p=0.0047) with low and high-risk patients experiencing 3-year cumulative incidences of transformation of 5.2% [95% CI 0.2%-10.2%] and 28.6% [95% CI 3.1%-54.0%] respectively, after adjusting for death as a competing risk. Notably, only 10 of 108 (9%) cases in the test set transformed to BP within 5-years and yet MPN13 still achieved significance in this cohort that had not previously seen the signature for training purposes.

## DISCUSSION

The need for accurate risk stratification for MF patients is paramount given the wide range of clinical outcomes experienced by these patients. To address this, current models for risk stratification use clinical features (i.e. DIPSS) (Passamonti et al. 2010) and more recently incorporate genomic mutations (i.e. MIPSS70) and/or cytogenetics (Guglielmelli et al. 2018; Grinfeld et al. 2018) to predict patient risk and tailor current treatment options. However, challenges in using these models remain. For example, acquiring mutation data can be cost-prohibitive and methods and informatic pipelines for acquiring and calling high molecular risk mutations are not yet standardized. Acquiring cytogenetic information from bone marrow samples is also challenging due to fibrotic marrows that are customary in MF patients.

Finally, while DIPSS is the most widely used risk stratification approach due to its low technical barrier, the vast majority of patients are classified as Intermediate risk (i.e. DIPSS-Intermediate-1 and DIPSS-Intermediate-2) where clinical decision making remains most unclear and where further resolution is warranted. Further, since none of these models consider the fundamental transcriptomic properties of the MF stem cells that drive disease outcomes, there remains an opportunity to complement and augment risk stratification approaches in MF. To address this, we used transcriptomic features from retrospective analysis of a published dataset of CD34+ MF stem cells (Psaila et al. 2020) to derive a short list of 1000 MF stem cell related genes on which to train prognostic models predictive of OS in MF patients from RNAseq data generated in PB-MNCs. This culminated in a final 24-gene expression signature (termed, MPN24) predictive of OS and a 13-gene subsignature (termed, MPN13) predictive of blastic transformation in MF. Importantly, MPN24 held prognostic value in multivariable analysis with DIPSS, MIPSS70 and Genomic-PRS indicating it captures information unique from clinical or mutational features of disease. This results in the ability to generate synergistic models, exemplified by our 3-tier risk stratification model that integrates DIPSS with MPN24.

Some limitations in our study remain. First, while we validated MPN24 and MPN13 in test set patients that were left out of signature training, additional cohorts will be needed to test the robustness of the signatures. Second, lack of consistent cytogenetic data prevented analysis of DIPSS-plus and MIPSS70-plus models and limited the scope of features included in the Genomic-PRS analysis. Future cohorts with comprehensive cytogenetics may be needed to test the added value MPN24 might offer to models informed by cytogenetics data. Further, while we present our new 3-tier risk stratification approach that integrates DIPSS and MPN24 it is likely that prospective clinical trials will be required to determine how this additional information should inform routine clinical practice, especially as novel therapies for MF continue to emerge.

While we anticipate MPN24 and MPN13 to powerfully impact MF patient care, future work will be needed to ensure clinical implementation with wide scale uptake. This might include, for example, measuring MPN24 and MPN13 with low-cost alternatives to RNA sequencing, such as real-time PCR or NanoString-based approaches. Platforms with high scalability, low-cost and rapid turnaround time will be critical to informing patient care in real time at diagnosis but also potentially, at additional time points across the patient journey. For example, longitudinal monitoring may be used to track disease severity during MF or as early as antecedent PV and ET phases. Further, since MPN24 is derived from features defining heterogeneity among MF stem cells, we hypothesize that the genes captured in this signature may also function as a powerful biomarker for upfront patient selection in clinical trials or informing appropriate timing of HCT. Additional studies will be required to validate these potential applications and build towards more precision medicine approaches in MF that are informed by the molecular nature of the MF stem cells that drive disease.

## Data Availability

Data produced in the present study are available upon reasonable request to the authors.

## Support

Program project grant to VG from The Elizabeth and Tony Comper Foundation; VG is supported by Barbara Baker Chair Award for Leukemia and related disorders; JJFM is the recipient of the Canadian Institutes of Health Research Doctoral Award: Frederick Banting and Charles Best Canada Graduate Scholarships [FBD-170928].

## Conflicts of Interest

VG reports consultancy: AbbVie, Bristol Myers Squibb–Celgene, Novartis, Pfizer, Daiichi Sankyo, GSK, Incyte; data safety or advisory board participation: AbbVie, Bristol Myers Squibb–Celgene, GSK, Incyte; honoraria: Bristol Myers Squibb–Celgene, Novartis; research support: Novartis, AbbVie. JED has licensing agreements with trillium Therapeutics and Pfizer for SIRP-a and has a sponsored research agreement with BMS.

## Notes

### Competing Interest Statement

VG reports consultancy with AbbVie, Bristol Myers Squibb/Celgene, Novartis, Pfizer, Daiichi Sankyo, GSK, Incyte; data safety or advisory board participation with AbbVie, Bristol Myers Squibb/Celgene, GSK, Incyte; honoraria from Bristol Myers Squibb/Celgene, Novartis; research support from Novartis, AbbVie. JED has licensing agreements with trillium Therapeutics and Pfizer for SIRP-a and has a sponsored research agreement with BMS.

### Author Declarations

Research Ethics Board of University Health Network gave ethical approval of this work (REB #17-5601).

